# Active follow-up of *Plasmodium vivax* radical treatment in a mobile and hard-to-reach population in the Amazon: results from the CUREMA project

**DOI:** 10.64898/2026.05.08.26352538

**Authors:** Alice Sanna, Yann Lambert, Lorraine Plessis, Carlotta Carboni, Teddy Bardon, Marion Petit-Sinturel, Irene Jimeno-Maroto, Muriel Galindo, Hedley Cairo, Josiane Müeller, Jane Bordalo Miller, Stephen Vreden, Martha Suarez-Mutis, Maylis Douine

## Abstract

**Background:** Evidence from real-world implementation of follow-up and pharmacovigilance for *Plasmodium vivax* treatment in population scale interventions targeting asymptomatic individuals remains limited, especially among highly mobile, hard-to-reach groups. This study describes the follow-up strategy used in the CUREMA project and assesses its implementation, as well as safety, tolerability, and adherence outcomes.

**Methods:** The CUREMA project implemented a PvTDA (*P. vivax* targeted drug administration) intervention in cross-border Amazonian settings, consisting in a full course treatment with chloroquine and primaquine (7 days) or tafenoquine (single dose), after screening its main contraindications (including semiquantitative G6PD testing); its target population was represented by artisanal and small scale migrant miners evaluated as at risk for asymptomatic *P. vivax* carriage. Treatment follow-up was mandatory for PvTDA participants and combined in-person visits or telephone interviews, and/or a tailored smartphone application, to be compatible with target population high mobility. Planned follow-up visits were scheduled 2, 5, and 14 days after treatment initiation. Univariate analyses described app uptake, follow-up completion, adverse events and treatment declared adherence; multivariable regression models explored factors associated with these outcomes.

**Results:** Among 294 participants who received PvTDA, 269 (91.5%) configured the smartphone application, 100 (34.0%) selected telephone follow-up and/or 44 (15.0%) selected in-person follow-up. Overall, 210/294 participants (71.4%) completed at least one follow-up questionnaire, and 81/294 (27.6%) completed the three follow-up visits At least one adverse event was recorded in 49/294 participants (16.7%); events were generally mild to moderate, and no serious adverse event was identified. The declared adherence to the 7-days chloroquine and primaquine treatment was 79.2% [95% CI 72.5-86.4].

**Conclusions:** Active follow-up of *P. vivax* radical treatment was feasible in a highly mobile, remote population when multiple complementary tools were combined. This supports pharmacovigilance and safe implementation of radical cure strategies in similarly challenging settings.

## Introduction

The global efforts toward *Plasmodium vivax* elimination have made significant progress with recent advances in diagnostic and therapeutic tools [1, 2]. The introduction of tafenoquine—a single-dose 8-aminoquinoline—and rapid G6PD testing has expanded treatment options [3, 4], while serological screening integration is considered to detect asymptomatic reservoirs of *Plasmodium vivax* [5, 6]. The World Health Organization (WHO) advises programmes to adopt targeted and reasoned strategies to accelerate malaria elimination, such as population-wide test-and-treat (TaT) approaches for active case detection, targeted drug administration (TDA), and mass drug administration (MDA), utilizing epidemiological or biological (molecular, serological) criteria to detect and treat asymptomatic infections, which constitute a major reservoir of the disease [1]. However, these population-wide interventions raise critical ethical and operational challenges, particularly regarding the risk-benefit balance of administering antimalarials to asymptomatic individuals. Beyond clinical efficacy, the tolerability, safety, and acceptability of these treatments, especially in real-world settings, remain poorly documented, yet they are pivotal to ensuring community uptake and program success.

A key concern is the paucity of real-world data on antimalarial adverse effects (AEs) outside controlled clinical trials. While older regimens (e.g., primaquine) are generally considered well-tolerated, several studies have retrospectively documented the occurrence of serious adverse events associated with these treatments (in the absence of G6PD testing), and the underestimation of adverse effects due to the limitations of prospective follow-up of patients undergoing treatment and the lack of intentional collection of this data [7, 8]. Moreover, most safety and tolerability data stem from symptomatic infections: the tolerability profile in asymptomatic individuals—who may perceive fewer direct benefits from treatment—remains understudied. Ethically, documenting the safety of population-level interventions like *P. vivax* TDA (PvTDA) is essential to justify their risks, particularly when targeting mobile or hard-to-reach populations with limited healthcare access.

Additionally, adherence to multi-dose regimens (e.g., primaquine) or the irreversible hemolytic risks of single dose tafenoquine pose distinct challenges. Though tafenoquine’s single-dose regimen may improve compliance, its lack of reversibility demand rigorous post-treatment monitoring [9], which is often considered logistically infeasible in remote settings.

The CUREMA project (2022-2025) implemented and evaluated a multicomponent intervention pursuing malaria elimination among artisanal and small-scale gold miners in the Guiana Shield, a mostly migrant population characterized by high mobility, geographic isolation, and low health literacy [10–12]. Among other activities, the CUREMA project included the delivery of *P. vivax* treatment to individuals considered at risk to be asymptomatic carriers. The treatment follow-up, safety monitoring, and adherence assessment was one of the objective of the project [12, 13].

To address these gaps, this paper describes: (i) the feasibility of antimalarial treatment follow-up in a hard-to-reach population settings; (ii) the observed safety profile of PvTDA in this context, including participant-reported adverse events; and (iii) self-reported adherence to primaquine and chloroquine, and associated factors.

## Materials and methods

The CUREMA project is a multicentric international study including a complex intervention targeting malaria and its evaluation. It has been fully described in Sanna et al. [10]. Data was collected at the time of the intervention itself, and by qualitative and quantitative studies realized before and after the intervention [10].

### The inclusion in the intervention

The target population of the intervention consisted of adults engaged in artisanal and small-scale gold mining in the region (inclusion criteria in Table 1). This group, consisting mainly of migrants of Brazilian origin, is characterized by significant commuting between the mining sites, located deep in the forest, and the logistical support bases situated in key transit towns and along the rivers that serve as access routes to the gold-mining areas in the forest.

**Table 1:** Inclusion criteria for the overall intervention and eligibility criteria for the PvTDA module.

| Inclusion criteria - Intervention | Eligibility Criteria - PvTDA |  |
| --- | --- | --- |
| • Be 18 years of age or older<br>• Weight over 35 Kg<br>• Agree to participate in the study<br>• Have a current involvement in gold mining activities (having been to the mining areas in the last year or planning to enter the mining areas in the following two weeks), regardless of country<br>• Not having symptoms of malaria at the time of the inclusion visit | • Wish to receive the module.<br>• Epidemiological criteria in favor of a current asymptomatic carriage of *P. vivax* (blood stage or liver stage). At least one of the following conditions:<br>• have a history of clinical malaria during the past 12 months.<br>• OR having a life-long malaria history AND have stayed for at least 1 week during the last 12 months in an area with extensive *P. vivax* transmission | • Accept to participate in active follow-up during the 14 days following the start of treatment.<br>• No current pregnancy (declared or rapid urine test positive) nor breastfeeding.<br>• Hemoglobinemia above 9 g/dL<br>• G6PD activity above 6 IU/gHb.<br>• Not having received a unique dose of tafenoquine within the last 3 months nor a full course of primaquine within the last month.<br>• No hypersensitivity nor known contraindication to chloroquine, primaquine or tafenoquine.<br>• No history of severe mental health disorder<br>• Not having a positive malaria rapid diagnostic the day of the inclusion nor currently receiving an anti-malarial treatment |

The intervention activities were carried out by facilitators trained by the project, who had a profile of community health workers; inclusions were conducted between March 2023 and December 2024 in key locations for cross-border mobility of gold miners on the rivers marking the borders between French Guiana and Brazil and Suriname as well as in the Surinamese capital Paramaribo (Figure 1).

**Figure 1:**
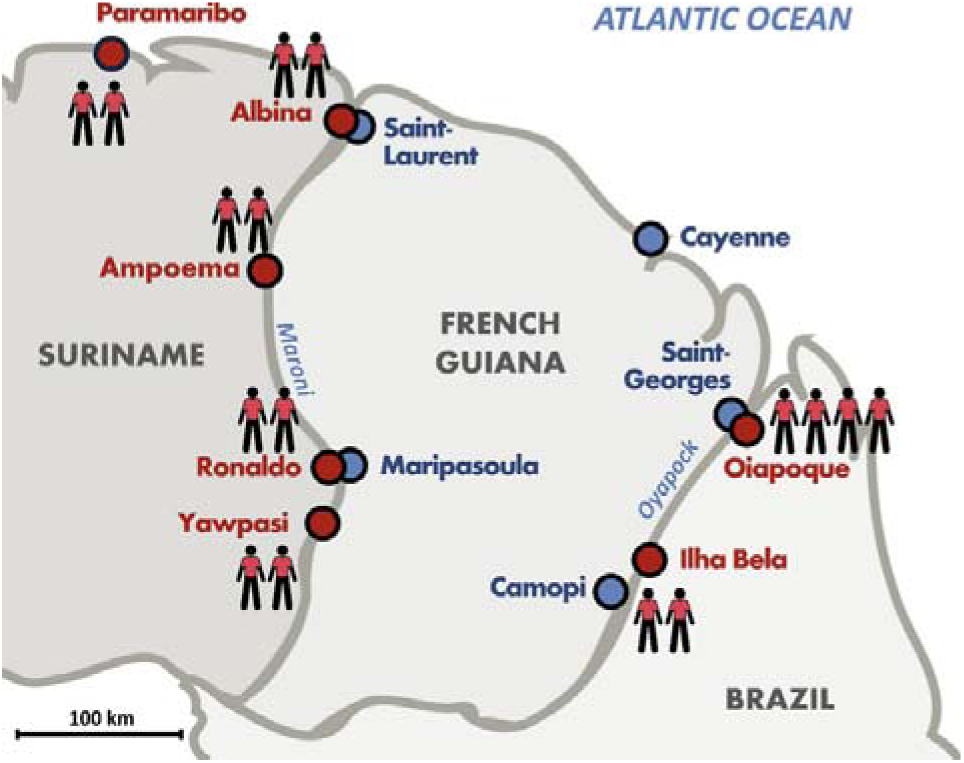
Map of the Guiana Shield representing the inclusion sites of the CUREMA intervention (red dots) with the respective sizes of local study’s field teams.

The intervention consisted of communication, information, and health education activities targeting malaria, as well as two modules offered to participants: i/ the free distribution of malaria self-test and self-treatment kits accompanied by training for participants (Malakit module) [11, 14]; ii/ treatment of asymptomatic individuals considered at risk of asymptomatic carriage of *Plasmodium vivax* (hypnozoites and/or circulating parasites at low concentrations), defined as PvTDA. PvTDA consisted in the administration of 3-days chloroquine and an 8-aminoquinoline (0.5mg/kg/day primaquine for 7 days or 300 mg tafenoquine in a single-dose) according to the weight categories shown in Table 2. The selection of PvTDA patients was based on voluntary participation and eligibility criteria: having a clinical and epidemiological history consistent with recent exposure to and infection with *P. vivax*, and no known contraindications (Table 1). Treatment was supervised for the first dose and self-administered for subsequent doses.

**Table 2:** Categories for weight adjusted posology for chloroquine (3 days) and primaquine (7 days) in the PvTDA module.

| Weight (kg) | Cholorquine tablets<br>(150 mg of<br>chloroquine base<br>equivalent) | Primaquine tablets<br>(15 mg) | OR Primaquine<br>tablets (7,5<br>mg) |
| --- | --- | --- | --- |
| 35-49 | 3, 3, 1 | 1 | 2 |
| 50-69 | 4, 4, 2 | 2 | 4 |
| 70-89 | 5, 5, 3 | 3 | 5 |
| >=90 | 6, 6, 3 | 3 | 6 |

The medication was provided along with verbal instructions regarding how to take it (including taking it with meals to reduce gastrointestinal side effects), follow-up procedures, the main potential side effects, and instructions to contact the study team (medical investigators) if any side effects occurred. At the time of enrollment, the medication was provided to the patient in a bag containing the daily doses grouped into small zip-top bags, each labeled with the respective day of treatment; an illustrated leaflet accompanied the bag (Figure 2).

**Figure 2:**
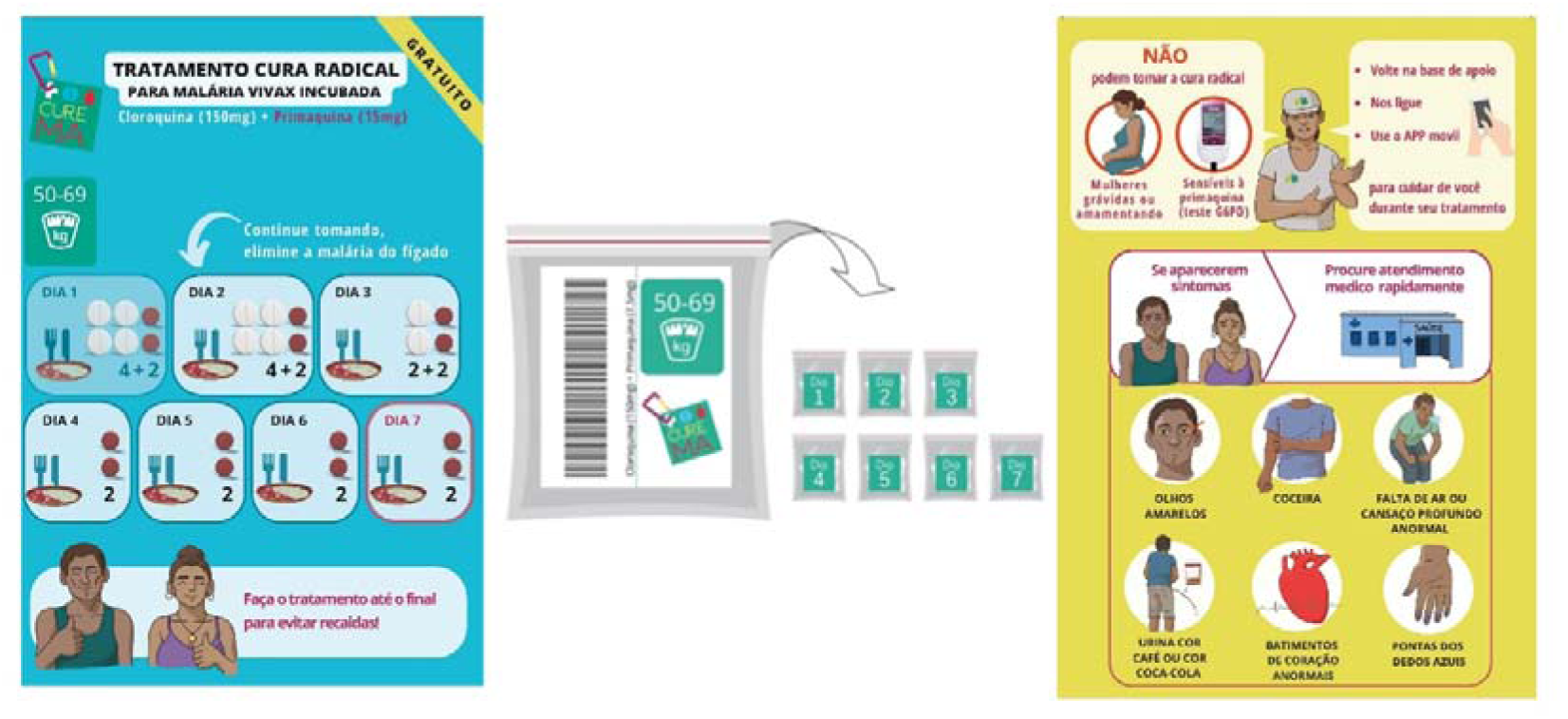
Illustration of an example of the PvTDA packaging and information leaflet (for the weight category 50-69 kg), with instructions on how to take the treatment (front, in light blue) and on main contraindications and adverse events (back, in yellow).

**Figure 3:**
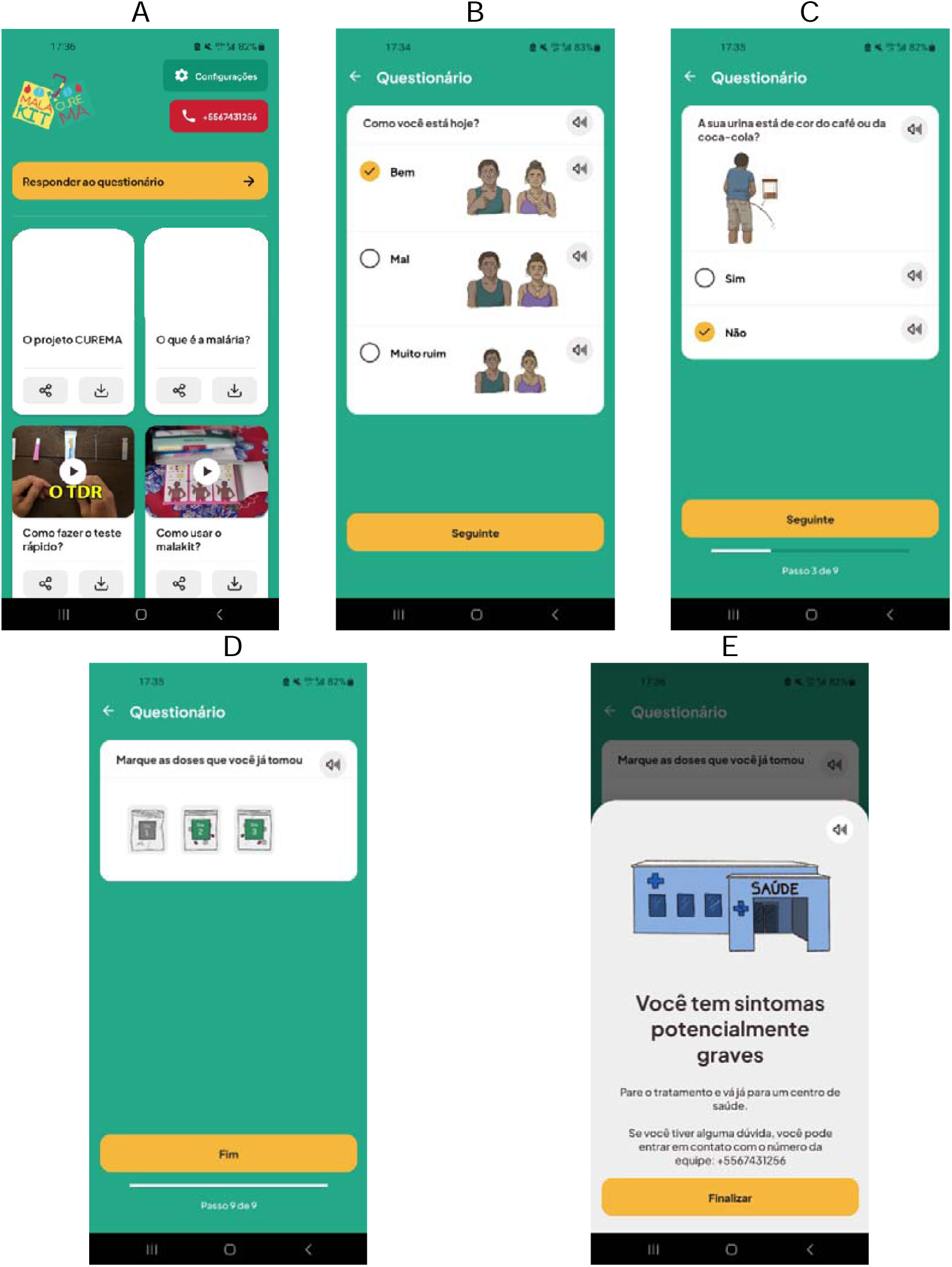
Screenshots of the final version of the smartphone application of the CUREMA project: A) initial screen, B) opening question of the questionnaire (“How are you today?”), C) one of the questions regarding the symptoms being assessed, D) question regarding adherence, E) message displayed in the event of warning symptoms.

The introduction of tafenoquine was only possible in Brazil from mid-August 2024 in the context of procurement’s obstacles.

### Data collection

Inclusion was determined and guided by data collected through a questionnaire that explored sociodemographic, occupational, and mobility variables, recent history of malaria exposure, as well as eligibility criteria for the intervention modules. To reduce the risk of iatrogenic harm due to dosing errors, the PvTDA bags were color-coded according to weight category; information regarding the weight category was also included in the barcode associated with each bag (which also contained information regarding the drug batch), which had to be scanned during the inclusion questionnaire, recording bag’s information in the participant’s data and triggering an alert if there was a mismatch between the participant’s weight and the category of the bag provided.

The data collection system was centered on ODK Collect [15], an open-source mobile data collection application installed on Android tablets at each inclusion site, working offline. One module supported all study questionnaires, and a second ODK Collect module was used to securely back up participant identification data from paper forms at the end of each day. These identification lists were encrypted and transmitted to a separate secondary ODK Cloud server, intentionally isolated from the primary server containing questionnaire data. The electronic identification backup supported pharmacovigilance activities by allowing rapid verification of participant information during adverse event investigations.

### PvTDA follow-up

#### Follow-up windows

Active follow-up was carried out for all participants during three visits on the following dates (considering the start of treatment as day 0): day 2 (window from day 2 to day 4), day 5 (5-13), and day 14 (14-19). Unplanned visits could be recorded before or after these periods in the event of unsolicited contact from a participant.

#### Follow-up methods

Participants could choose one or more follow-up methods: i/ to return in person to the inclusion sites and answer a face-to-face questionnaire from the project facilitators; ii/ to be contacted by telephone to receive the same questionnaire from the facilitators; and or iii/ to complete a simplified version of the questionnaire on an *ad hoc* smartphone application.

The questionnaires completed by the facilitators, as well as unanswered calls, were entered in real time on an electronic form implemented in the ODK collect follow-up questionnaires on the project tablets. Facilitators were also instructed to immediately report any adverse events they were aware of to the project coordination team.

The smartphone application was a software compatible with Android phones developed for the project by the company Appsolute SAS. The app was not available on public download platforms, in order to control its distribution and use in this pilot phase. To allow the smartphone app download in the participants phones despite the connectivity challenges—given that many inclusion sites lacked consistent internet access—each site was equipped with a Raspberry Pi device, generating a local Wi-Fi network (without internet access). When connected to the network and using their smartphone’s browser, the participant was automatically redirected to a webpage offering to download and install the app thanks to an apk file. This system was intentionally designed for simplicity, allowing facilitators to integrate it seamlessly into their daily workflows. The project’s application, installed on participants’ smartphones, contained informational videos and, for those receiving PVTDA, a customized follow-up module. The application module designed to monitor PvTDA was activated and configured at the time of inclusion with key information about the participant, allowing the questions to be personalized (type of treatment, weight, etc.) and an anonymity code to be assigned to link them to the participant’s identity. Designed for users with low literacy and limited access to eyecare, the application allowed for the configuration of voice reading of the questions as well as the choice of increased font sizes. The questions were accompanied by static or animated illustrations of the symptoms explored, validated through iterative consultation with community members. The completed questionnaires were presented on the theoretical follow-up dates via pop-up notifications and displayed daily until a response was given or a pre-set deadline was reached. If the participant had not answered the questionnaire by the deadline, an empty form was automatically generated and sent to the server to track the application’s activity and connectivity. At the participant’s discretion, it was also possible to set up daily reminders to take the treatment, accompanied by a simple question (“How are you today?”) which would bring up the follow-up questionnaire if the participant reported feeling unwell. In the event of warning signs (participant reporting feeling “very unwell” or presenting at least one symptom that could be associated with an AESI (adverse event of special interest), an alert message was displayed on the mobile phone screen instructing the participant to suspend treatment and seek care; the cell phone number of the team that enrolled the participant was also displayed. The data produced by the questionnaire (empty, partially completed, or fully completed) were stored locally in the cell phone’s memory and then transmitted to the server as soon as an internet connection was available, on mobile network or satellite Starlink access, increasingly available in gold mining areas [16].

Monitoring of the follow-up of participants who had received PvTDA was routinely carried out by the project coordination team, aggregating the data entered by the facilitators and obtained by the smartphone application, with reminders sent to the facilitators in the event of incomplete follow-up. The tafenoquine roll-out (Brazilian inclusion sites, August to December 2024) the follow-up was strengthened, systematically requiring its realization by facilitators (thus avoiding follow-up exclusively via the app).

Other potential sources of adverse events reporting during the project were the health-care facilities of the region, informed about the study, as well as the community itself, through its solidarity mechanism.

#### Outcomes explored

The follow-up questionnaires, in their complete version (completed by facilitators) and simplified version (application), explored symptoms potentially associated with AESI identified by the study protocol: acute hemolytic anemia, methemoglobinemia, allergic reactions, and heart rhythm disorders. The following symptoms were therefore actively explored: dyspnea, feeling unwell, altered heart rhythm, paleness, skin and mucous membrane jaundice, cyanosis of the extremities, dark urine, pruritus, and skin rash.

In addition, the questionnaire began by exploring the participant’s perceived general health and offered the possibility of reporting any other symptoms as well as any seeking of healthcare after the start of treatment.

Serious adverse events were classified in accordance with the following definition: unexpected events resulting in death, life-threatening condition, hospitalization or prolonged hospitalization, probable occurrence of permanent functional impairment.

Adherence was assessed by asking participants whether they were still taking their medication or if it had been prematurely stopped, and how many doses they had taken up to the time of the questionnaire. In the smartphone app, only the question about the number of doses taken was asked through graphical representation of the numbered treatment zip-top pockets.

#### Management of adverse event reported

Reported adverse events were investigated by telephone by one of the study’s medical investigators, either directly with the participant (preferred option) or with the facilitators who had communicated with the participant if the symptoms were very mild and already resolved or the participant could not be reached. The purpose of this investigation was to identify signs of potential severity, facilitate access to care if deemed necessary, and better characterize the event. The symptoms described in the questionnaires were, if necessary, reclassified (if the responses entered did not correspond to the symptoms described), and a causality analysis (probable/improbable) was performed by the investigator based on the timing and characteristics of the event described. The data collected were then centralized in descriptive spreadsheets to enable internal monitoring of adverse events and to be reported quarterly to the project’s DSMB (data and safety monitoring board) and the sponsor’s pharmacovigilance center.

If a serious adverse event was detected, it was immediately notified to the study sponsor, the ethics committee, and the competent pharmacovigilance authorities. In these cases, a double causality analysis was also planned (by the investigator and by the pharmacovigilance centre), as well as prospective follow-up of the patient.

### Other data sources

Results from pre- and post-intervention quantitative surveys (ORPAL 3, conducted in the last quarter of 2022, and ORPAL 4, conducted between December 2024 and April 2025) are also presented regarding contextual data on access to mobile phones and to internet at gold mining sites. Data were also collected by the project’s facilitators during enrollments carried out at the cross-border gold miners’ resting sites; the participants were adults involved in gold-mining, having left a French Guiana mining site since a week or less.

### Data analysis

Quantitative data are presented in aggregate form by figures and proportions, by medians and interquartile ranges, or by mean and standard deviation, as appropriate. Univariate and multivariate analyses were performed using logistic regression models to identify factors associated with participation in follow-up (defined as the presence of at least one completed follow-up questionnaire, by any means and on any date) and with presenting an adverse event (reporting one or more symptoms considered compatible in their nature and timing with the treatment under study). Treatment adherence (defined as not reporting that treatment had been prematurely stopped and a number of doses taken corresponding to the number of days since the start of treatment) is analyzed by presenting early treatment discontinuations using a Kaplan-Meier curve and analyzed using a Cox model.

Statistical analyses were performed using R software (version 4.4.0) and R-Studio (version RStudio 2025.09.2)

## Results

Overall, 2,002 inclusions were realized in the CUREMA project intervention and among them, 294 led to a PvTDA delivery.

During the ORPAL 3 and ORPAL 4 surveys were included respectively 540 and 487 persons, ( 212 and 197 in Brazil, and 328 and 290 in Suriname).

### Feasibility and acceptability of PvTDA follow-up among the study population

#### Access to mobile phone and internet connection in the study population (pre- and post-intervention surveys)

During the ORPAL 3 study, more than half of the gold miners reported having access to the internet at their last gold mining site (n=327, 60.6%), and for 64.5% of them (n=214), the connection required payment. Half of participants in this survey (n=268, 49.6%) reported having connected to the internet with their cell phone at least once in the previous 14 days.

Two years later, according to the ORPAL 4 data, a larger part of the goldminers working in French Guiana had access to internet in the last mining-site (86%, n=413), and 94%, (n=451) declared owning a smartphone. Among the persons owing a smartphone, 93% (n=420) reported having an Android operating system.

#### Smartphone app download among the participants in the CUREMA intervention

During the 2,002 inclusions in the CUREMA project, in 1,417 (70.8%) the project app was successfully installed on the participant’s smartphone. Among the most common reasons for not installing the app (multiple reasons were possible), 194 participants (9.7%) stated that they did not have a phone with them at the time of enrolment, 206 (10.3%) stated that they did not own a phone, 62 (3.1%) owned a phone with an iOS operating system, and only 35 (1.7%) expressed a refusal to install the app.

#### Quantitative insights on follow-up acceptability for CUREMA participants

In the 1,025 inclusions in which participants refused participate in the PvTDA module, 19 (1.3%) stated that they declined due to the constraints imposed by follow-up.

Of the 570 inclusions in which the participant chose to participate in the PvTDA module, 6 had to be excluded because of the follow-up: 5 refused it afterward and 1 had no cell-phone and was not able to participate to in person visits.

Among those who opted to receive the PvTDA module, the median expected length of stay at the inclusion site (before returning to the mines or leaving for another place) was 5 days (Q1-Q3 = 2-7 days).

### Follow-up implementation among the participants in the CUREMA PvTDA intervention

#### Characteristics of participants having received a PvTDA intervention

As illustrated in Table 3, the majority of the 294 inclusions with PvTDA delivery were carried out in Suriname (n=175, 59.5%); 147 of these inclusions were made in urban areas (50%) in Oiapoque, Albina, or Paramaribo, 15 (5.1%) directly at the gold mining site in Lourenço (Amapá state, Brazil), and 132 (40.5%) at remote logistical rear bases (Ilha Bela, Ampuma, Ronaldo or Yawpassi). The median age of these participants was 42 (ranging from 19 to 75), and the M: F sex ratio was 2.46; 16.3% (n=48) of them had no formal education, and 50% (n=147) had only completed primary school. Half of the participants were directly involved in gold extraction (n=136, 46.3%), the others worked in operational support roles; the median length of time spent in gold mining was 10 years (Q1-Q3 = 6-20 years). The majority were at the inclusion site to prepare for an upcoming trip to a gold mining site (92, 31.3%), to make purchases (79, 26.9%) and/or to rest (80, 27.2%). The most common destination immediately after was a gold mining site in French Guiana (243, 82.7%) or in Brazil (22, 7.5%). Regarding malaria history, 140 (47.6%) reported having had malaria symptoms in the past year, 101 (34%) reported having received a biological diagnosis of malaria (with or without symptoms) during the same period; 125 (42.5%) reported having already received primaquine as part of radical treatment during their lifetime.

**Table 3:**
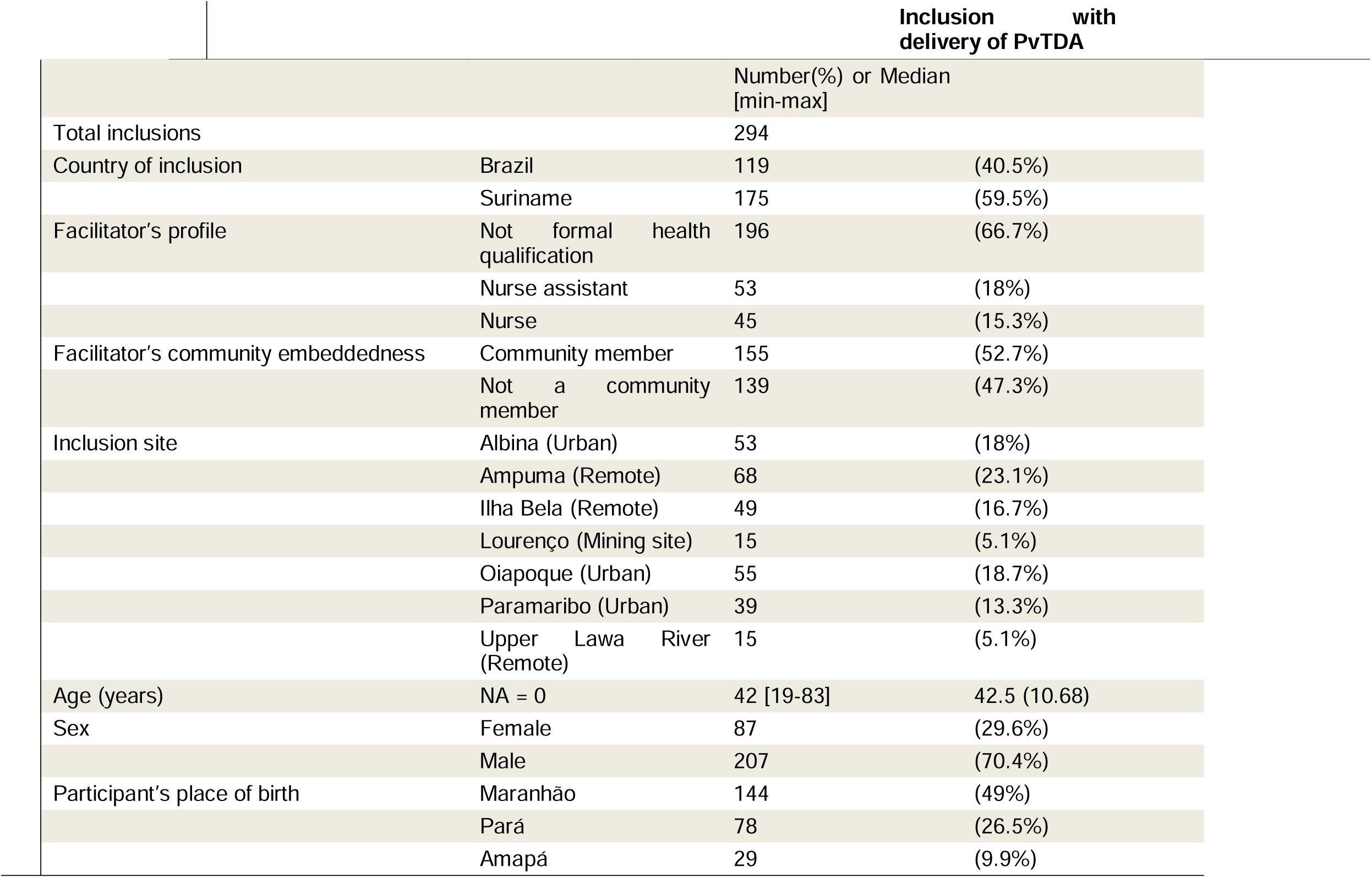
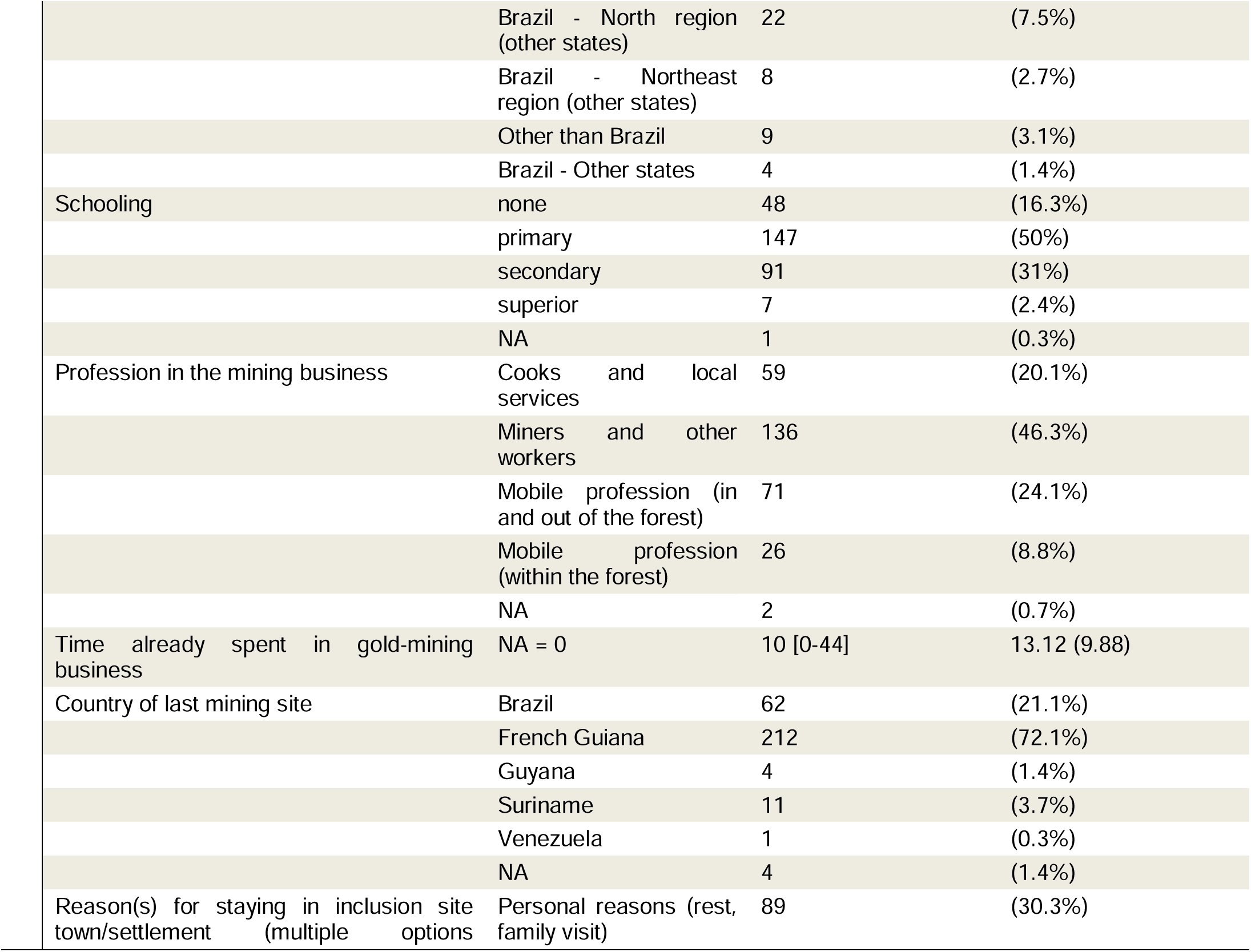
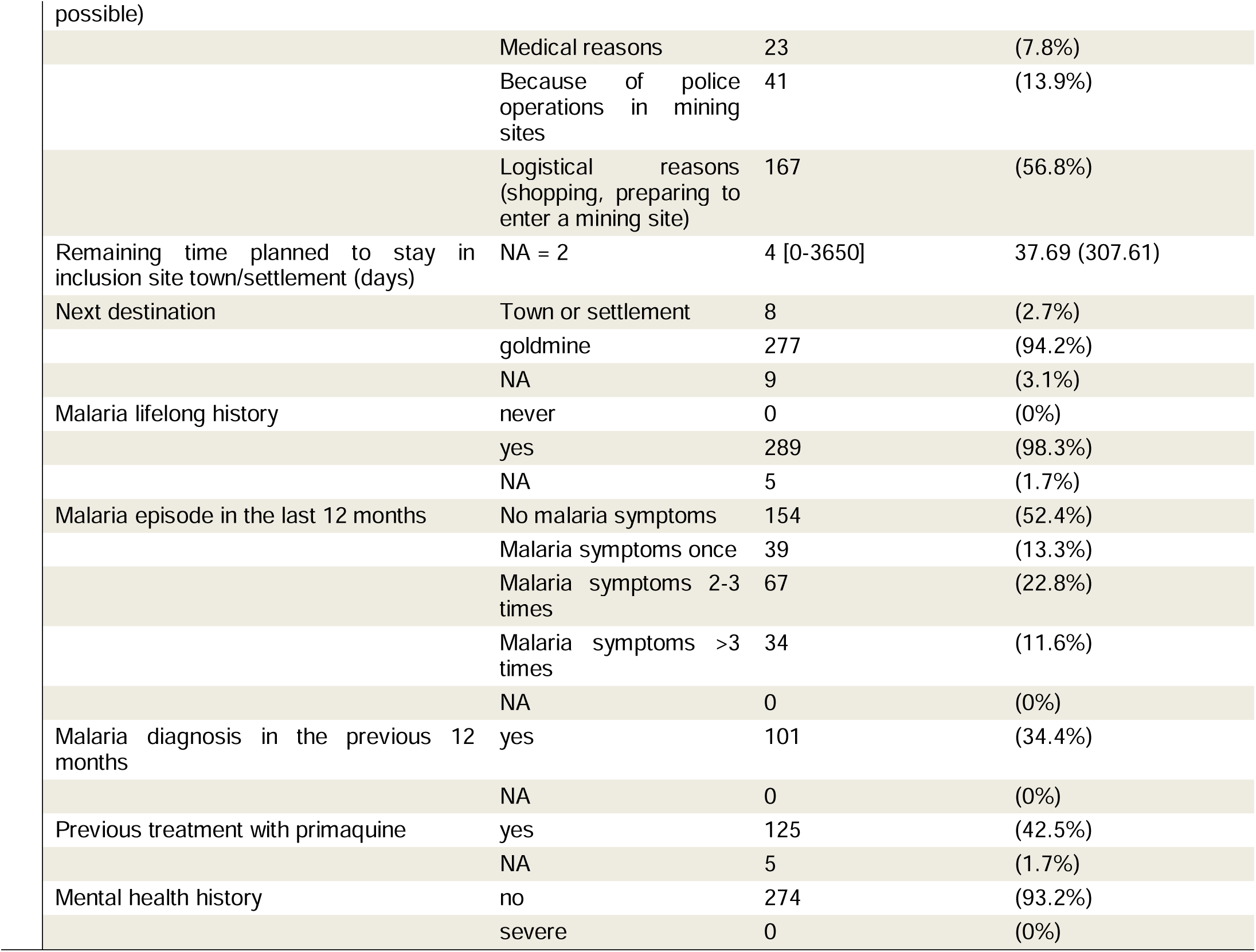
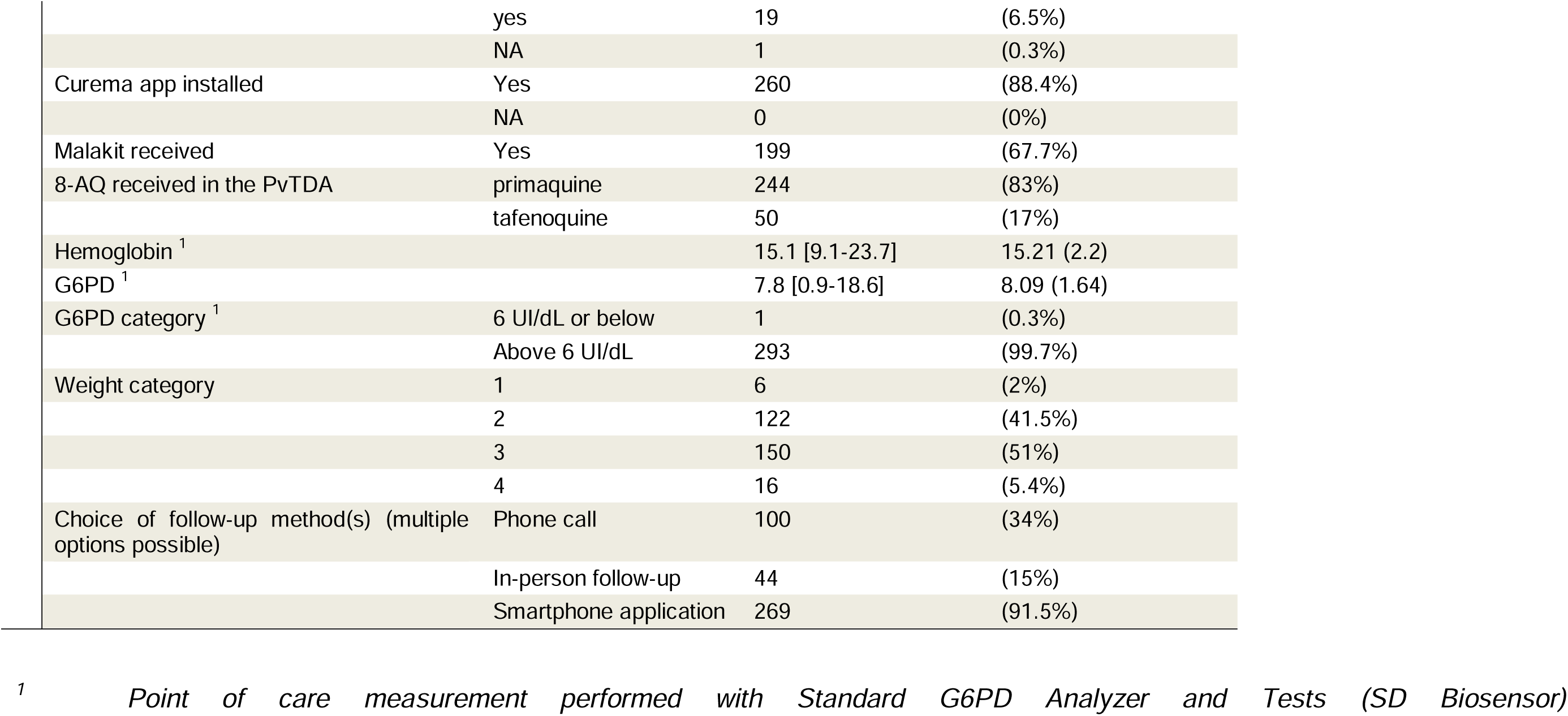
Characteristics of the inclusions in the CUREMA intervention involving a PvTDA module delivery.

|  |  | Inclusion with delivery of PvTDA |  |
| --- | --- | --- | --- |
|  |  | Number(%) or Median [min-max] |  |
| Total inclusions |  | 294 |  |
| Country of inclusion | Brazil | 119 | (40.5%) |
|  | Suriname | 175 | (59.5%) |
| Facilitator's profile | Not formal health qualification | 196 | (66.7%) |
|  | Nurse assistant | 53 | (18%) |
|  | Nurse | 45 | (15.3%) |
| Facilitator's community embeddedness | Community member | 155 | (52.7%) |
|  | Not a community member | 139 | (47.3%) |
| Inclusion site | Albina (Urban) | 53 | (18%) |
|  | Ampuma (Remote) | 68 | (23.1%) |
|  | Ilha Bela (Remote) | 49 | (16.7%) |
|  | Lourenço (Mining site) | 15 | (5.1%) |
|  | Oiapoque (Urban) | 55 | (18.7%) |
|  | Paramaribo (Urban) | 39 | (13.3%) |
|  | Upper Lawa River (Remote) | 15 | (5.1%) |
| Age (years) | NA = 0 | 42 [19-83] | 42.5 (10.68) |
| Sex | Female | 87 | (29.6%) |
|  | Male | 207 | (70.4%) |
| Participant's place of birth | Maranhão | 144 | (49%) |
|  | Pará | 78 | (26.5%) |
|  | Amapá | 29 | (9.9%) |
|  | Brazil - North region (other states) | 22 | (7.5%) |
|  | Brazil - Northeast region (other states) | 8 | (2.7%) |
|  | Other than Brazil | 9 | (3.1%) |
|  | Brazil - Other states | 4 | (1.4%) |
| Schooling | none | 48 | (16.3%) |
|  | primary | 147 | (50%) |
|  | secondary | 91 | (31%) |
|  | superior | 7 | (2.4%) |
|  | NA | 1 | (0.3%) |
| Profession in the mining business | Cooks and local services | 59 | (20.1%) |
|  | Miners and other workers | 136 | (46.3%) |
|  | Mobile profession (in and out of the forest) | 71 | (24.1%) |
|  | Mobile profession (within the forest) | 26 | (8.8%) |
|  | NA | 2 | (0.7%) |
| Time already spent in gold-mining business | NA = 0 | 10 [0-44] | 13.12 (9.88) |
| Country of last mining site | Brazil | 62 | (21.1%) |
|  | French Guiana | 212 | (72.1%) |
|  | Guyana | 4 | (1.4%) |
|  | Suriname | 11 | (3.7%) |
|  | Venezuela | 1 | (0.3%) |
|  | NA | 4 | (1.4%) |
| Reason(s) for staying in inclusion site town/settlement (multiple options) | Personal reasons (rest, family visit) | 89 | (30.3%) |
|  | Medical reasons | 23 | (7.8%) |
|  | Because of police operations in mining sites | 41 | (13.9%) |
|  | Logistical reasons (shopping, preparing to enter a mining site) | 167 | (56.8%) |
| Remaining time planned to stay in inclusion site town/settlement (days) | NA = 2 | 4 [0-3650] | 37.69 (307.61) |
| Next destination | Town or settlement | 8 | (2.7%) |
|  | goldmine | 277 | (94.2%) |
|  | NA | 9 | (3.1%) |
| Malaria lifelong history | never | 0 | (0%) |
|  | yes | 289 | (98.3%) |
|  | NA | 5 | (1.7%) |
| Malaria episode in the last 12 months | No malaria symptoms | 154 | (52.4%) |
|  | Malaria symptoms once | 39 | (13.3%) |
|  | Malaria symptoms 2-3 times | 67 | (22.8%) |
|  | Malaria symptoms >3 times | 34 | (11.6%) |
|  | NA | 0 | (0%) |
| Malaria diagnosis in the previous 12 months | yes | 101 | (34.4%) |
|  | NA | 0 | (0%) |
| Previous treatment with primaquine | yes | 125 | (42.5%) |
|  | NA | 5 | (1.7%) |
| Mental health history | no | 274 | (93.2%) |
|  | severe | 0 | (0%) |
|  | yes | 19 | (6.5%) |
|  | NA | 1 | (0.3%) |
| Curema app installed | Yes | 260 | (88.4%) |
|  | NA | 0 | (0%) |
| Malakit received | Yes | 199 | (67.7%) |
| 8-AQ received in the PVTDA | primaquine | 244 | (83%) |
|  | tafenoquine | 50 | (17%) |
| Hemoglobin <sup>1</sup> |  | 15.1 [9.1-23.7] | 15.21 (2.2) |
| G6PD <sup>1</sup> |  | 7.8 [0.9-18.6] | 8.09 (1.64) |
| G6PD category <sup>1</sup> | 6 UI/dL or below | 1 | (0.3%) |
|  | Above 6 UI/dL | 293 | (99.7%) |
| Weight category | 1 | 6 | (2%) |
|  | 2 | 122 | (41.5%) |
|  | 3 | 150 | (51%) |
|  | 4 | 16 | (5.4%) |
| Choice of follow-up method(s) (multiple options possible) | Phone call | 100 | (34%) |
|  | In-person follow-up | 44 | (15%) |
|  | Smartphone application | 269 | (91.5%) |

<sup>1</sup> Point of care measurement performed with Standard G6PD Analyzer and Tests (SD Biosensor)

Two-third also received a malakit (199, 67.7%); the others were unable to benefit from this service essentially due to a temporary stock shortage.

Regarding the PvTDA module, 244 participants (83%) received treatment with primaquine and chloroquine, while the remaining 50 (17%) received treatment with tafenoquine and chloroquine. At inclusion, the chosen follow-up options were : by phone (100,34%), in person (44, 15%) or through the app (269,91.5%).

### Follow-up effectiveness and associated factors

Two third of the participants responded to at least one follow-up questionnaire (210, 71.4%). The latest answer was by day 21, with a median response time to the first questionnaire of 3 days (Q1-Q3 = 1-4 days) - except one participant who responded to a questionnaire on day 207 after starting treatment Phone calls were the most commonly used follow-up method (124, 42.2%), closely followed by the smartphone application (114, 38.8%), while face-to-face visits were less frequent (50, 17.0%). A comparison between the type(s) of follow-up chosen and the type(s) follow-up actually carried out is presented in Table 4.

**Table 4:** Comparison between the follow-up methods chosen at inclusion and those actually used during the follow-up.

|  | Choice of in person follow-up |  | Choice of Phone call follow-up |  | Choice of smartphone app follow-up |  |
| --- | --- | --- | --- | --- | --- | --- |
|  | No | Yes | No | Yes | No | Yes |
| No visit in person | 230 (92%) | 14 (31.9%) | 166 (85.6%) | 78 (78%) | 16 (64%) | 228 (84.8%) |
| At least 1 visit in person | 20 (8%) | 30 (68.2%) | 28 (14.4%) | 22 (22%) | 9 (36%) | 41 (15.2%) |
| No phone questionnaire | 148 (59.2%) | 22 (50%) | 140 (72.2%) | 30 (30%) | 11 (44%) | 159 (59.1%) |
| At least 1 phone questionnaire | 102 (40.8%) | 22 (50%) | 54 (27.8%) | 70 (70%) | 14 (56%) | 110 (40.9%) |
| No app questionnaire | 160 (64%) | 20 (45.5%) | 106 (54.6%) | 74 (74%) | 25 (100%) | 155 (57.6%) |
| At least 1 app questionnaire | 90 (36%) | 24 (54.5%) | 88 (45.4%) | 26 (26%) | 0 (0%) | 114 (42.4%) |

**Table 5:**
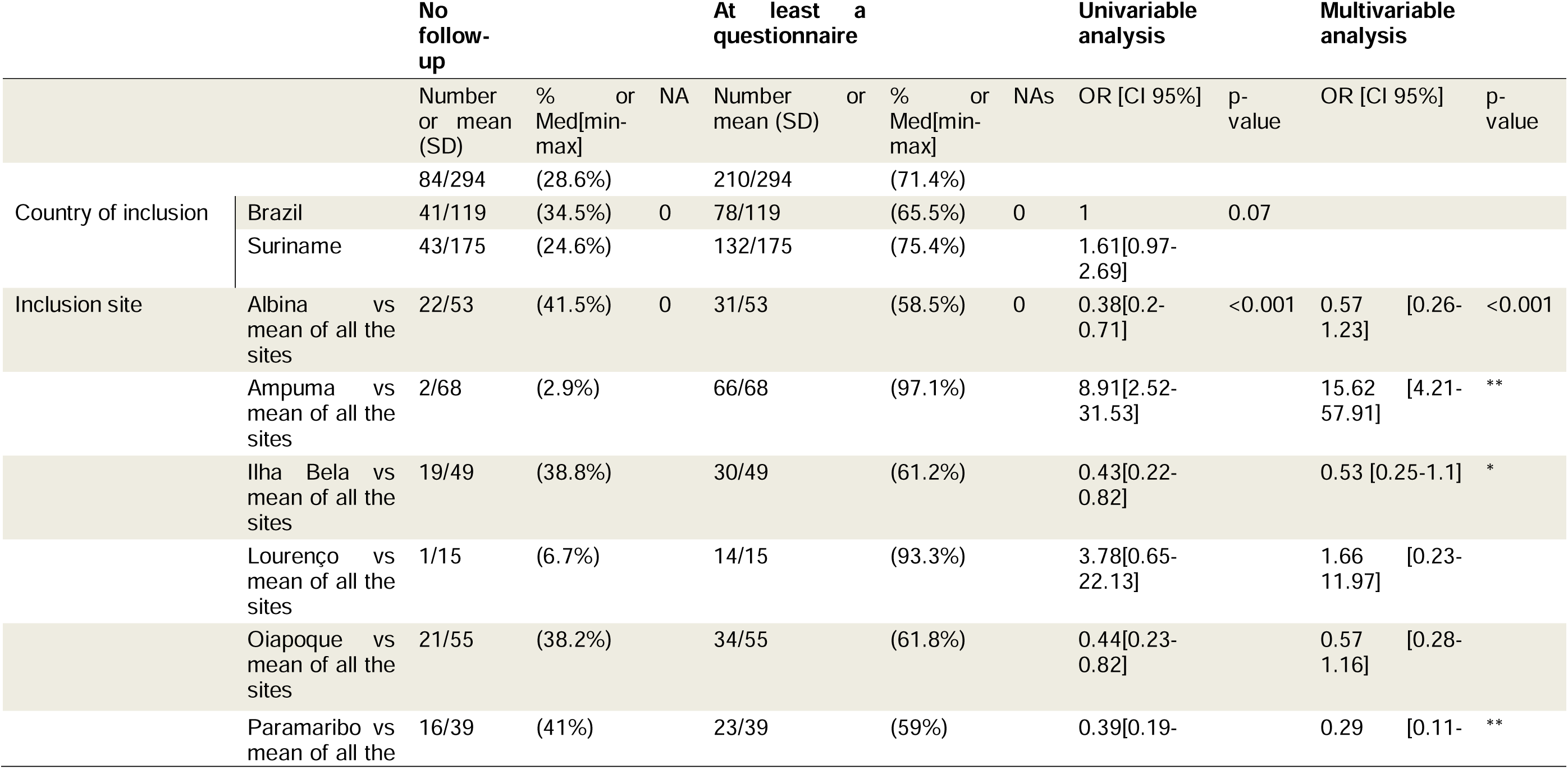
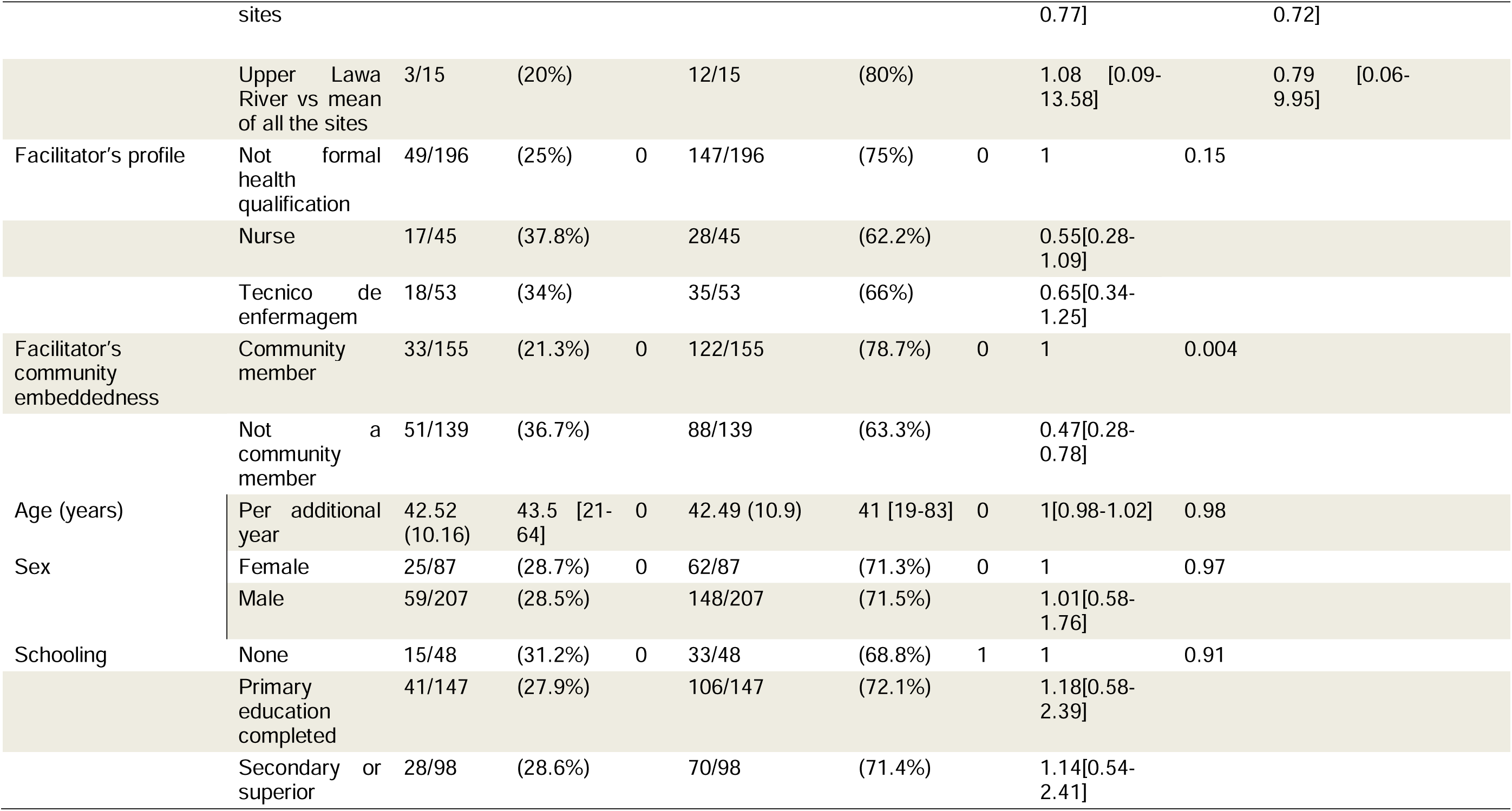
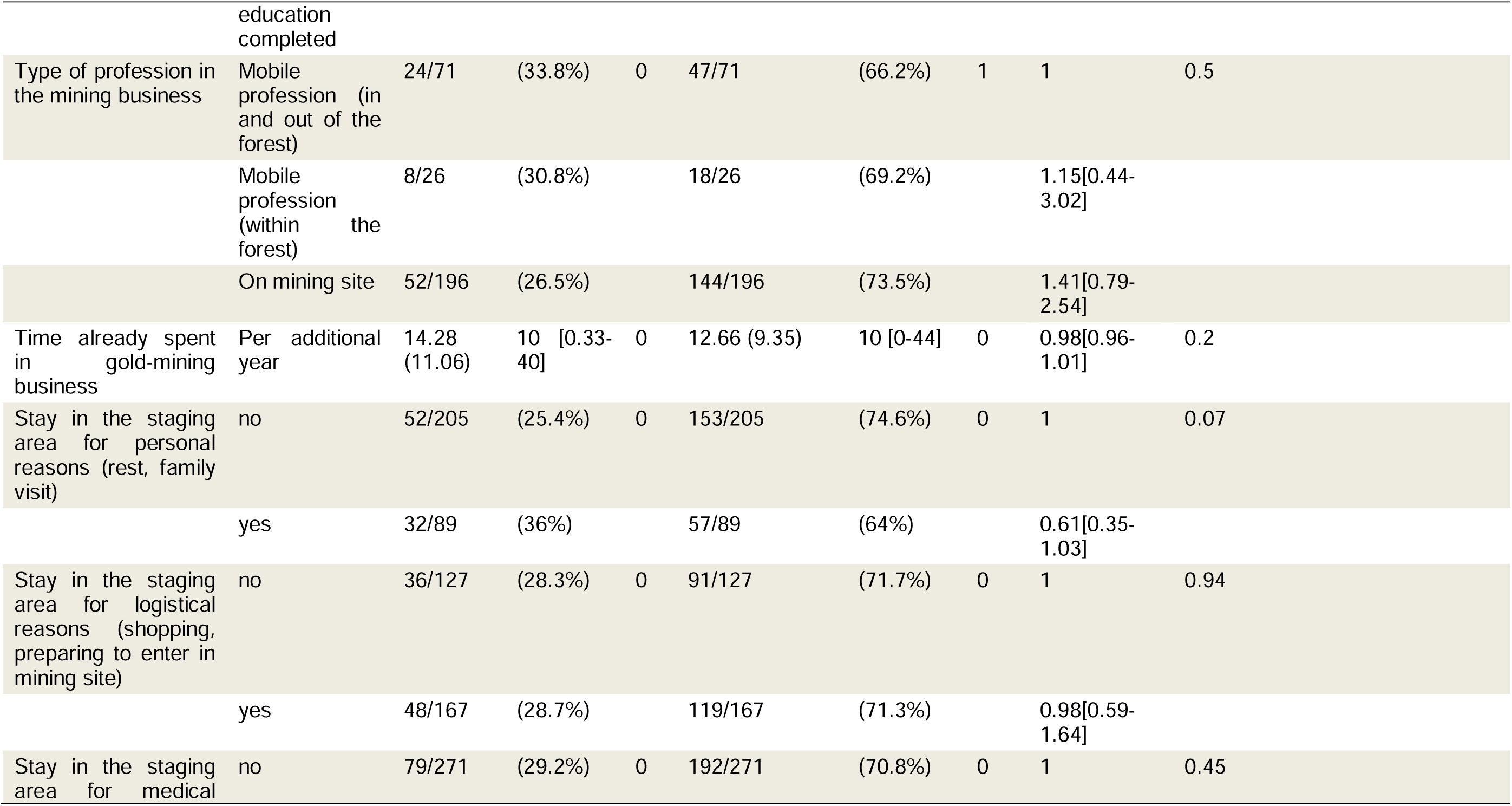
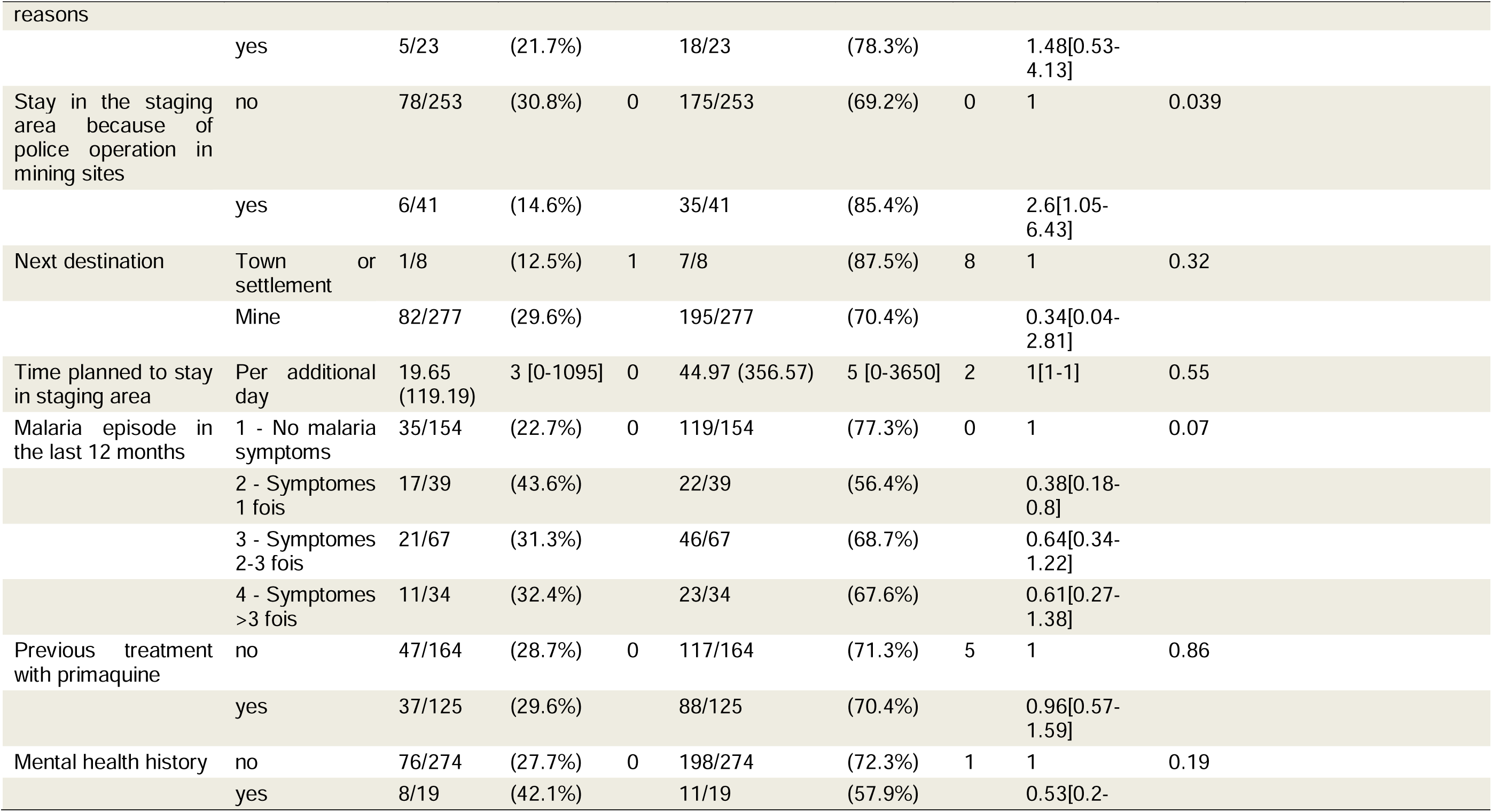
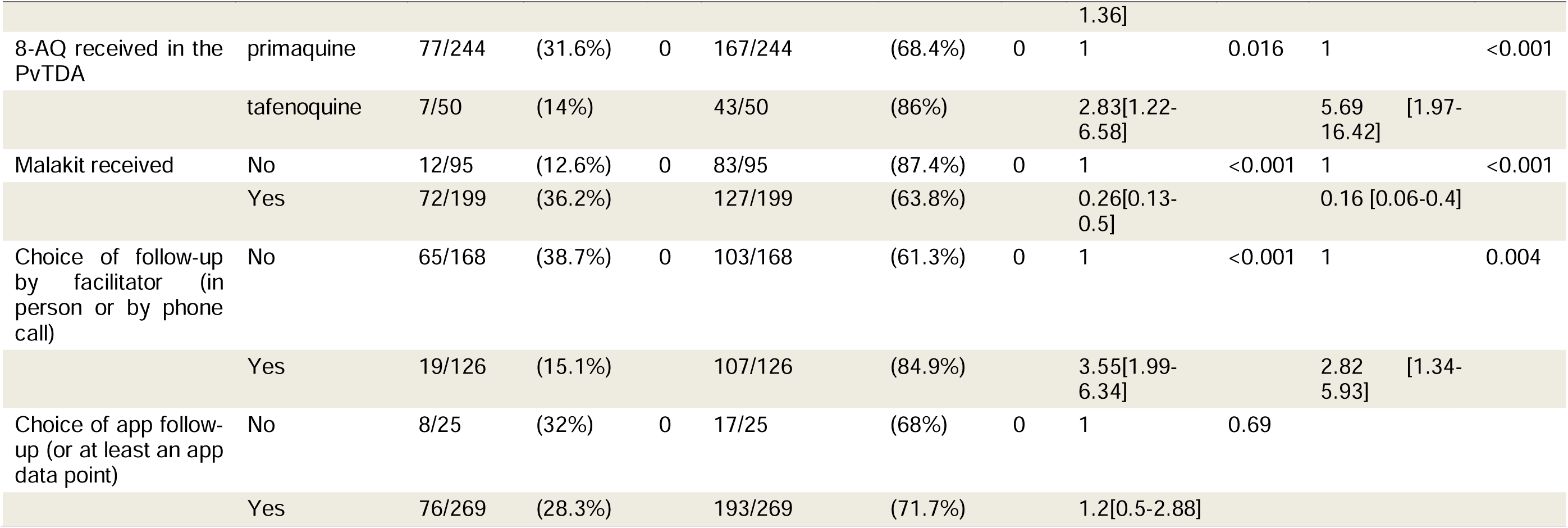
Characteristics of PvTDA participants according to the effectiveness of their follow-up. (SD = standard deviation; Med = median; min = minimum; max = maximum; NA = not available; OR = Odds ratio; CI = confidence interval)

Among the 147 participants who completed at least one questionnaire (in person or by telephone call) with a facilitator, the median number of questionnaires completed was 2 (Q1-Q3 = 1-3). Among the 114 participants who responded to at least one questionnaire in the smartphone app, the median number of forms completed was 5 (Q1-Q3 = 1-8).

As illustrated in Figure 4: Figures and proportion of follow-up visits completed bu PvTDA participants. more than a half of the participants answered at least one questionnaire in the two critical follow-up windows (between day 2 and 4 and between day 5 and 13).Overall 81 participants (27.6%) had a follow-up questionnaire completed for all the three planned inclusion visit in their specific windows.

**Figure 4:**
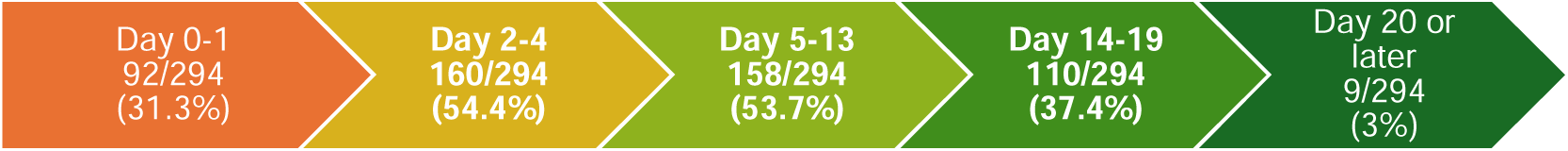
Figures and proportion of follow-up visits completed bu PvTDA participants.

A total of 628 non-empty questionnaires were sent via the smartphone app. Of these, 468 (74.5%) were sent to the server instantly. When delayed (160 questionnaires), the median time between form submission and server receipt was 5 hours (Q1-Q3 = 20 minutes – 35.5 hours), with a 95 percentile of 2.6 days and a maximum delay of 165 days.

The factors associated with follow-up participation are described in Table 3. In multivariable analysis, the factors favorably associated were certain sites of inclusion, particularly Ampuma (OR 15.62, 95% CI [4.21-57.91]); having received tafenoquine (OR 5.69, 95% CI [1.97-16.42]) and having accepted at least one follow-up method lead by the facilitators (phone call and/or in-person visit) (OR 2.82, 95% CI [1.34-5.93]). Having been able to receive Malakit was unfavorably associated with effective follow-up (OR 0.16, 95% CI [0.06-0.4]).

### Adverse events and safety of the treatment

Forty-nine participants (16.7%) recorded at least one adverse event during treatment: 48 during the follow-up and one reported by the community network without direct contact with him being possible. In addition, two participants mistakenly sent questionnaires in the smartphone app reporting adverse events (wrong handling).

Of these 51 alerts received (49 AEs and 2 wrong handlings): 29 (56.9%) were investigated by telephone by one of the investigating physicians; 7 (13.7%) were considered unnecessary (very mild symptoms that had already resolved), and 15 (29.4%) investigations were not possible (the participant was not reachable by call on the phone contact informed at inclusion and had not replied to the messages sent by the investigator on messaging app).

The evaluation of these events, based on the information available in the questionnaires and, where applicable, in the telephone investigation, identified symptoms probably related to the PvTDA treatment in 40 participants (39 of the 210 people with follow-up, 18.6%), for 7 (3.3%) participants symptoms probably unrelated to the treatment received, and for 2 (1%) participants symptoms for which it is not possible to take a position based on the available information. The latter participants will be considered, as a conservative measure, as having experienced adverse effects related to PvTDA in the subsequent analyses.

Thirty one adverse events (73.4%) were considered mild in intensity and 9 (21.4%) moderate in intensity. No serious adverse event was identified. Four participants seek care for their symptoms in an outpatient facility, and 3 were oriented by the investigator to a medical assessment but refused it; all of them reported symptoms resolution without further intervention. The median time of symptom onset was day 2 of treatment, with 29 (74.3%) participants reporting symptoms that appeared between Day 1 and Day 3. Details of the described symptoms are shown in Table 6.

**Table 6:** Main symptoms reported by PvTDA participants.

| Symptom | Frequency | Proportion (n=211) |
| --- | --- | --- |
| Dizziness | 15 | (7.1%) |
| Hearth rhythm change | 13 | (6.2%) |
| Nausea and vomiting | 8 | (3.8%) |
| Asthenia | 7 | (3.3%) |
| Cutaneous rash | 6 | (2.8%) |
| Ocular symptoms | 6 | (2.8%) |
| Itch | 5 | (2.4%) |
| Abdominal pain | 5 | (2.4%) |
| Diarrhea | 5 | (2.4%) |
| Headache | 4 | (1.9%) |
| Psychological symptoms | 4 | (1.9%) |
| Dyspnea | 3 | (1.4%) |
| Dark urine | 2 | (0.9%) |
| Pale skin | 1 | (0.5%) |

Other symptoms were also reported in the questionnaires and telephone interviews: productive cough, upper respiratory tract infection, episode of malaria, skin fungus, green coloration of urine.

Table 7 describes the characteristics of participants who were followed up according to whether they reported an adverse event. The only factors significantly associated at multivariable analysis with a lower risk of reporting of adverse events were the site of inclusion (particularly Ampuma (OR 0.21 [ CI95% 0.08-0.54]) and male gender (OR 0.29 [CI95% 0.13-0.62]).

**Table 7:**
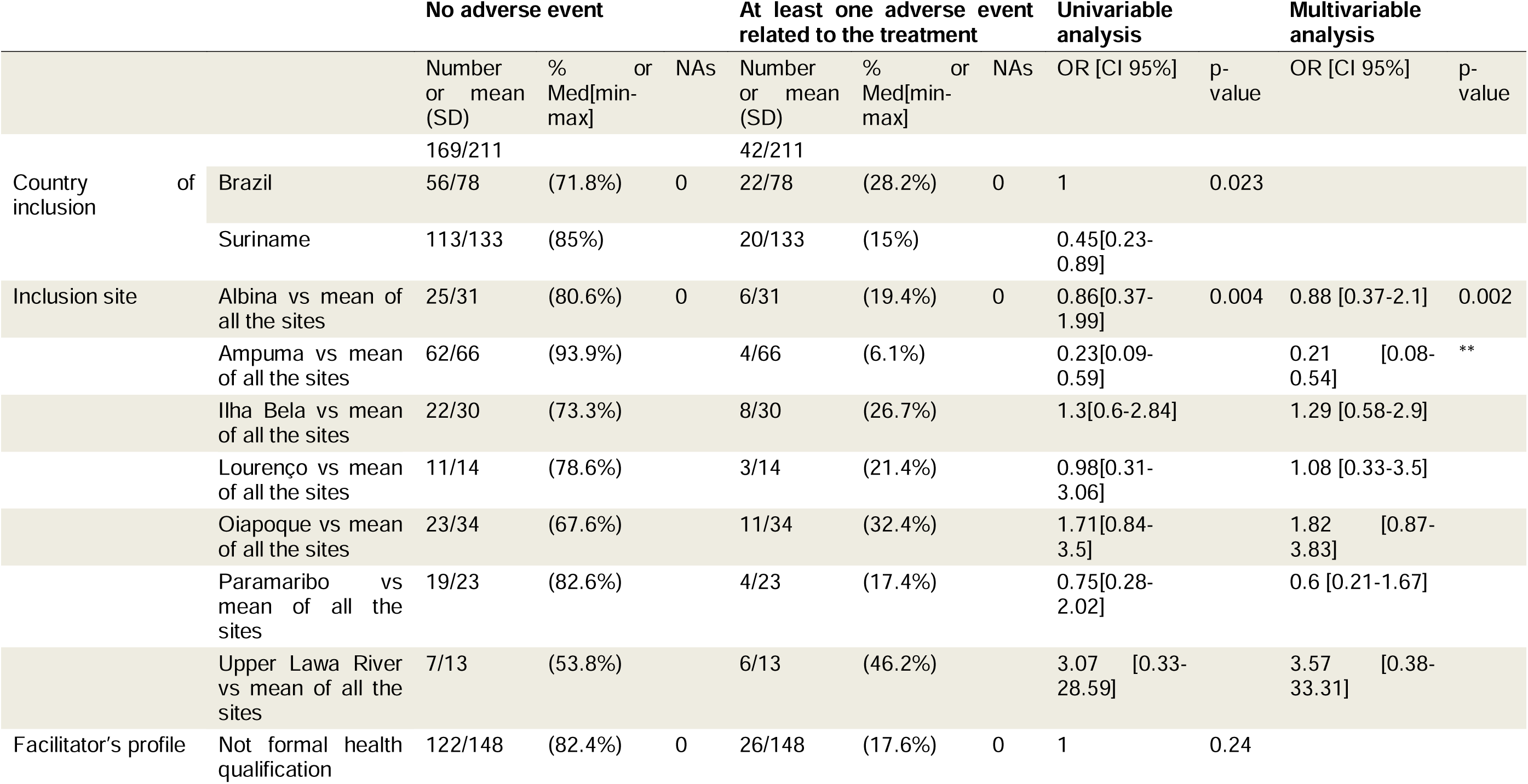
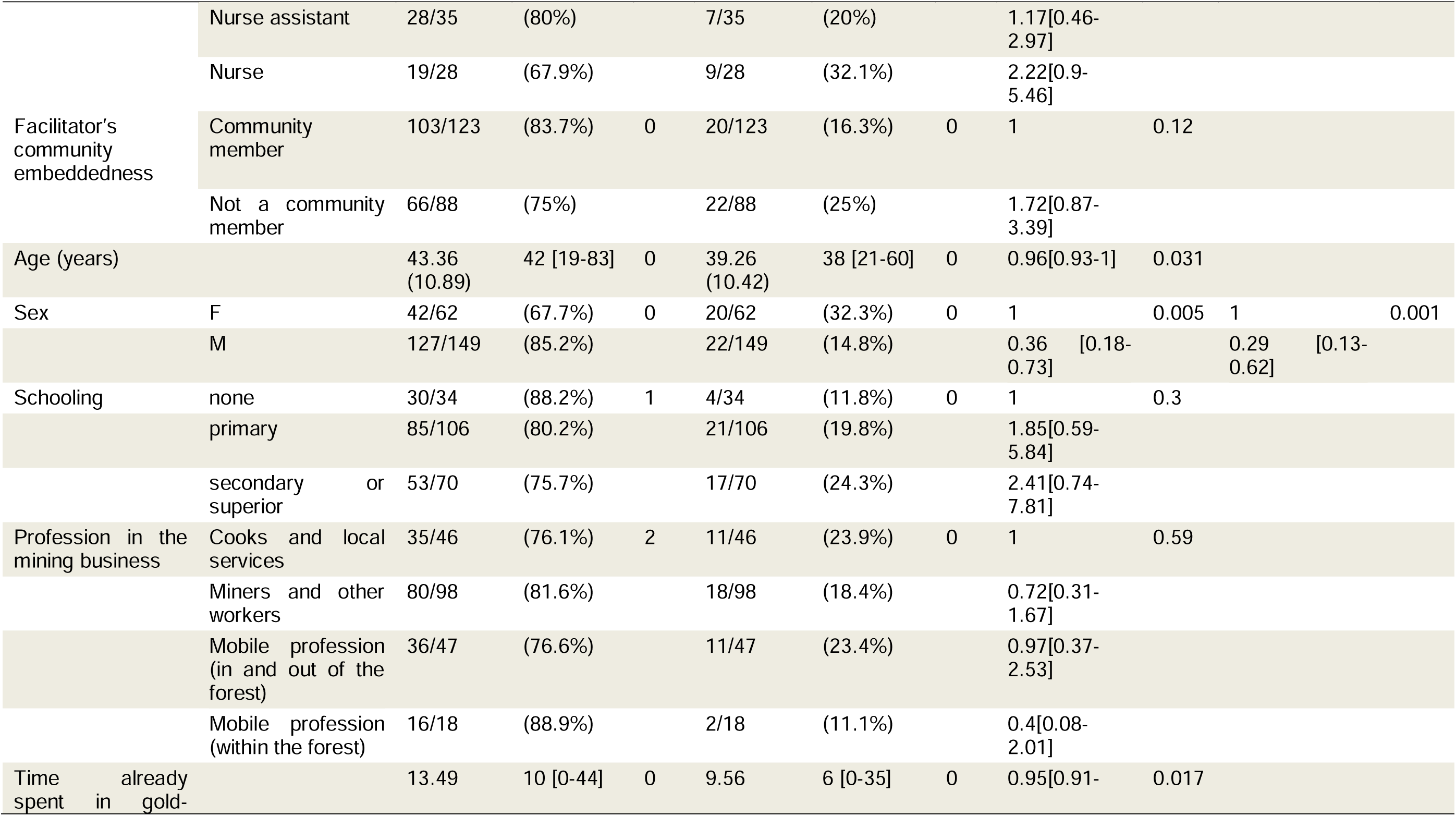
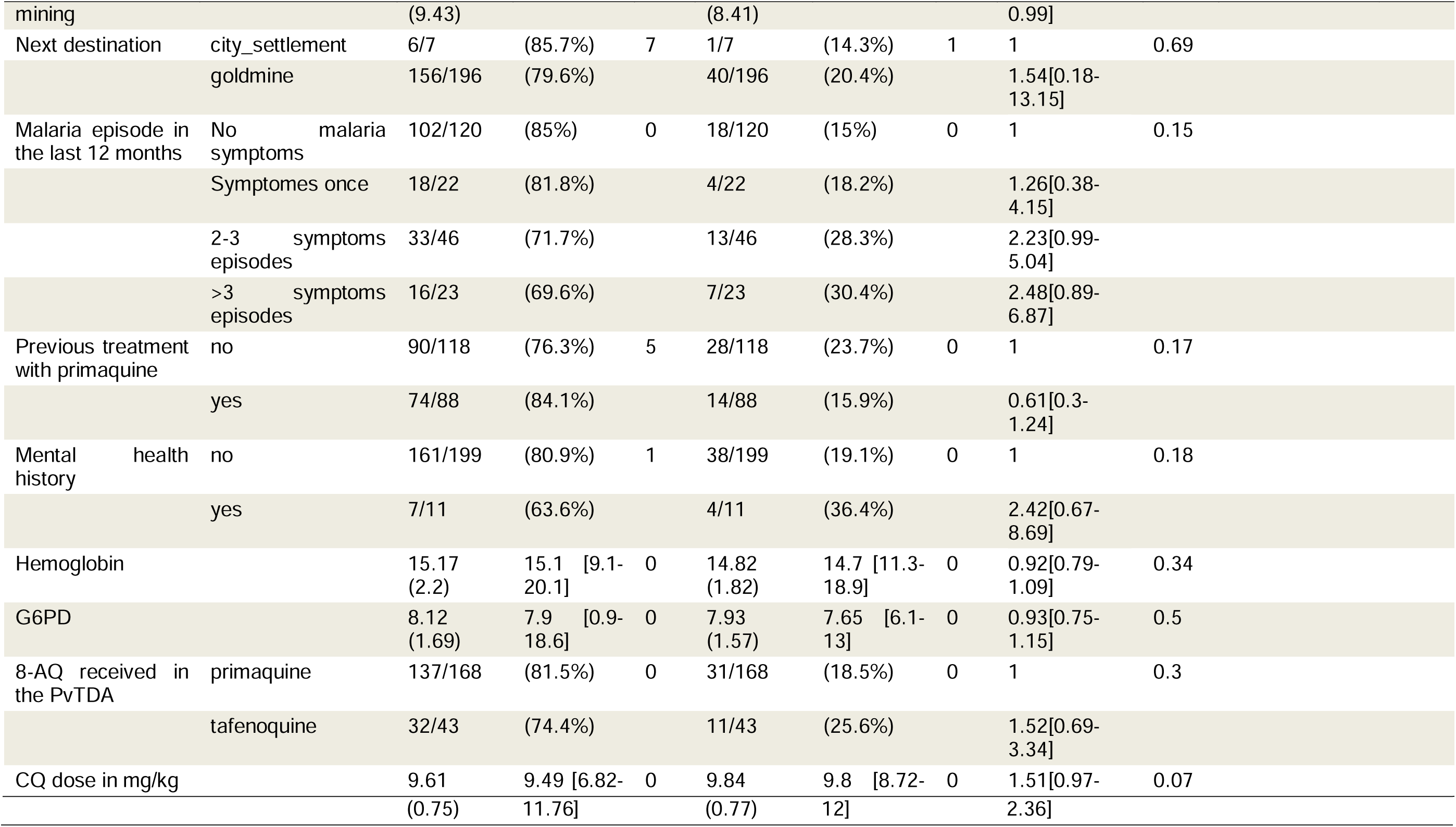
Factors associated with report of adverse event among participants in PvTDA with follow -up (N=211, French Guiana, Curema Proejct, 2023-2024). (SD = standard deviation; Med = median; min = minimum; max = maximum; NA = not available; OR = Odds ratio; CI = confidence interval)

A multivariable analysis realized on the subgroup of participants having received primaquine found similar results, and in addition identified a significant positive association between the daily dose of primaquine in mg/kg and the odds of reporting an adverse event (OR 2.25 [CI95% 1.07-4.73] per additional 0.1 mg/Kg/day) (Supplementary materials).

#### Community detection of adverse events and alert system

The lack of a stable community structure linked to gold mining and the fluidity of interpersonal relationships, with no community leader who could act as a stable point of contact for external actors, made it impossible to set up a formal community alert system for adverse events. Nevertheless, the community was the source of two reports of events, which reached the investigation team via the project facilitators: one regarding mental health symptoms occurred in a participant having received the PvTDA in Suriname, reported by the manager of his mining site to the facilitators; one report of probable an acute hemolysis occurred in a former participant who were excluded from PvTDA because of G6PD deficiency but later received primaquine in the context of malaria episode diagnosed by the health system teams in Brazil before the implantation of the G6PD test in their treatment protocol in 2024.

#### Other safety alerts

During the inclusions in the PvTDA module, the choice of treatment delivery (whether or not the treatment should be administered and at what dosage) was supported by the electronic questionnaire’s decision algorithm. A single incident was reported: a facilitator misinterpreted the result on the G6PD Analyzer device screen, entering in the electronic form 9 UI/dL instead of 0.9 UI/dL, which led to the incorrect administration of PvTDA to a participant who probably was G6PD deficient. This error, detected by systematic cross-checking of pictures showing G6PD results, did not result in any adverse events: the participant had completed his treatment without the onset of any symptoms.

### Treatment adherence

Reported adherence to CQ+PQ treatment at day 7 was estimated at 79.2% [95% CI 72.5-86.4] (Supplementary materials). For chloroquine and tafenoquine treatment, the first dose was observed (adherence to TQ treatment was thus 100%), the adherence at day 3 was 90.5% [95% CI 82-99.8%].

In the multivariate analysis, early treatment discontinuation (for primaquine) was negatively associated with male gender (HR 0.37, 95% CI [0.17-0.80]) and positively associated with reporting an adverse event (HR 4.04, 95% CI [1.91-8.58]).

## Discussion

This analysis shows, on the one hand, the feasibility of actively monitoring adverse events associated with antimalarial treatment, even in a highly mobile population living in isolated areas, provided that the methods used are adapted to the characteristics of this population. The success of the follow-up is linked primarily to implementation characteristics (team, concomitant activities, pressure from supervisors) rather than to patient characteristics. In terms of safety and tolerability, on the one hand, the intervention was shown to be safe; on the other hand, in the context of treating asymptomatic individuals, and when extensive information on the risks associated with treatment is provided in the research participation information sheet, relatively frequent adverse events are reported. This finding calls for close attention to the treatment context and the approach used to inform participants, in order to improve the acceptability of strategies designed to accelerate the elimination of malaria.

### Innovation and feasibility

A key point of the experience presented in this paper lies in the innovative proposal of remote active follow-up for *P. vivax* treatment among a highly mobile population in a remote cross-border setting: a combination of active outreach by community health workers with a smartphone application capable of collecting data and generating alerts offline. Implementing follow-up under these conditions was particularly ambitious given the high pace of miners’ mobility in the study area (median stay in the inclusion settlement, 4 days; median stay in the same gold-mining area, 3 months). Miners operate deep in the Amazon forest in areas that are difficult for health professionals to reach and that generally lack mobile phone coverage, aside from the recently introduced satellite internet hotspots provided by a growing proportion of mines owners [13, 16, 17]. Our system was conceived as a proof of concept for a *P. vivax* treatment follow-up approach, which has proven to be successful and could be adapted to similar, extremely challenging, settings.

Experience of interventions strengthening the follow-up of anti-malarial treatment in international literature presents interesting comparisons. In Thailand, an intervention piloted in the 2000s integrated web-based and mobile technologies for the notification, investigation, and follow-up of malaria cases by malaria staff, including CHWs, in remote border regions [18]. Day 7 follow-up participation improved significantly compared with baseline, even among migrants (from <10% to 94%), but it is important to nota that, in that success story, the migrant population appears to have consisted largely of rural laborers from neighboring countries with medium- to long-term mobility. In Brazil, two recent pilots have aimed to improve follow-up, particularly pharmacovigilance, for malaria treatments. The first intervention, Smart Safety Surveillance (3S) pharmacovigilance risk [19], was piloted in Amazonas state and included three activities for newly diagnosed *P. vivax* malaria cases: distribution of an illustrated leaflet with treatment instructions and explaining the main symptoms of acute hemolysis; enrollment in SMS reminders for treatment intake; and a follow-up phone call by malaria staff three days after treatment completion. Among patients who accepted SMS alerts (1,512/5,594; 27%), 70.2% answered the follow-up questionnaire, one of the main obstacles being poor mobile phone coverage in the study area. A second small-scale experience was conducted within the TRuST study, which evaluated tafenoquine implementation in the public health system [20]. WhatsApp messages were automatically generated and sent on day 7 and week 8 to a subset of patients who had received *P. vivax* treatment in Manaus and Porto Velho in order to collect pharmacovigilance data. Participants were asked whether they had experienced new symptoms or become pregnant and, if they answered yes, were contacted by pharmacovigilance staff: on day 7, 79% of patients replied to the follow-up message.

In the CUREMA intervention, overall, the proportion of PvTDA participants who were successfully followed up: 70% overall, with rates above 50% during the critical windows for acute hemolysis symptoms between day 2 and 6. These results appear satisfactory when compared those obtained in less challenging context described above, in settings with lower mobility and fewer connectivity constraints.

The combination of methods and technologies was a major strength, integrating face-to-face or telephone contact with trusted facilitators - more appropriate for reassurance and support when problems arose - with alternative strategies for data collection among participants who were more difficult to reach. Stakeholder and community consultation during the development and piloting of the follow-up approach [21] appears to have contributed importantly to its success, as reported in other studies [22]. Developing the smartphone application required a substantial investment of time and effort, including community consultation, co-construction, and iterative testing over nearly three years. The main challenge was designing an illustrated questionnaire that was truly accessible and easy to understand for the target population with low health literacy. The application created additional opportunities for follow-up, health promotion and information about malaria, but ultimately only a fraction of those who installed it actually transmitted data. It is impossible to determine whether non-transmission resulted from loss of the phone, uninstallation of the application before connectivity was restored, malfunction at the time of delayed transmission or unwillingness to use it finally. Follow-up through an internet-based messaging application such as WhatsApp, as described by Pereira and colleagues [20] for the TRuST experience, could potentially offer the offline data-collection component sought with the application, without adding complexity for users or the workload associated with designing and maintaining a tailored application.

The requirement to conduct follow-up did not appear as a barrier to participation in the study, as only a small number of participants refused or were classified as ineligible due to infeasible follow-up. However, this may still have been an undeclared reason for refusal, potentially linked to fear of being tracked and reported to the police through follow-up tools. Facilitators may also have preemptively excluded individuals who were unlikely to comply with follow-up requirements without documenting this information.

Notably, in multivariate analysis, the main factors associated with follow-up success reflect facilitators’ motivation and the implementation process: the site itself (team effect), tafenoquine use - as the new drug introduction was accompanied by a strict follow-up process imposed by the coordination team - and malakit distribution (availability of Malakit versus periods during which mediators focused exclusively on PvTDA-related tasks).

### Safety and Tolerability of the *Plasmodium vivax* treatment in PvTDA context

In this study, mild to moderate adverse events were reported in 23% of participants with a PvTDA follow-up, in which 18% were attributed to the study drugs by investigators. No serious adverse event was identified.

Comparing the proportion of observed adverse events with the literature is challenging. Most of the publications reporting data on the safety and tolerability of *P. vivax* treatment are issued from clinical trials or observational studies conducted among participants with symptomatic malaria, creating potential confusion between disease-related and drug-related symptoms: differences in symptom classification produce highly variable estimates [23–28]. In the context of treating asymptomatic individuals, the estimates also vary. Several mass treatment campaigns targeting *P. vivax* have been conducted since the 1950s by malaria elimination programs across the globe, which have treated millions of asymptomatic individuals with primaquine alone or in combination with anti-schizonticidal drugs; they are nevertheless poorly documented, do not systematically collect tolerability data, and report safety outcomes such as hospitalization or death for a reduced number of participants [29, 30]. Mass primaquine treatment conducted in recent years in South Korea and Myanmar with 15 mg of 14-days primaquine documented adverse events in 4 to 6% of participants, mainly headache, dizziness, and gastrointestinal symptoms [30, 31]. In Greece, MDA campaign using chloroquine plus 14 days of primaquine among 1,094 migrants led to 36.2% of participants reporting side effects during treatment, mainly dizziness, headache, and gastrointestinal disorders. In that study, most adverse events were attributed to chloroquine [32]. This pattern is consistent with the CUREMA experience (23%), in which most symptoms occurred between days 1 and 3 and resolved after completion or discontinuation of chloroquine.

Marked variability in side effects reported during antimalarial treatment has also been systematically described in a multi-country cluster-randomized trial comparing three anti-*P. vivax* treatment regimens: variability between sites was far greater than variability between study arms (high-dose primaquine, low-dose primaquine, and placebo) [23], despite standardized procedures and staff training. This heterogeneity may reflect both the posture of the professionals asking the questions and cultural factors shaping how symptoms are perceived and expressed. Such findings are consistent with the substantial heterogeneity in adverse-event reporting across inclusion sites observed in this article.

The nocebo effect, defined as adverse effects produced by expectations [33], is a well-documented phenomenon with a neurobiological basis and may help explain both the relatively high prevalence of reported side effects in this study and the heterogeneity in reporting. The informed-consent procedures, which included detailed explanations of the potential side effects of study drugs, may have contributed to expectation and increased the likelihood of symptom reporting [33]. Factors associated with nocebo effects include communication style (more or less reassuring), the patient-physician relationship and trust, drug characteristics (with stronger effects reported for generic and new drugs), and sociodemographic characteristics such as female sex, age, low socioeconomic status, and residence in remote areas [34]. This interpretation is consistent with the increased reporting of adverse events among female participants, which may reflect a combination of cultural, psychological, and pharmacokinetic factors.

The higher frequency of side effects may also have been related to the actual mg/kg dosage of primaquine. The study targeted an intermediate daily dose of 0.5 mg/kg/day, but the weight categories introduced some variation in the administered mg/kg dose. The distribution of weight categories - particularly category 2 - placed more women at the lower end of the category than men, resulting in slightly higher primaquine dosing in this group. However, the resulting doses remained within an intermediate range for primaquine (between 0.5 and 0.6 mg/kg/day), whereas more recent recommendations suggest increasing the dose to as much as 1 mg/kg [2, 24]. No overdose was therefore detected, although dose variation may have contributed in part to lower tolerability, particularly among asymptomatic individuals.

Among the main limitations about safety and tolerability description, the findings rely essentially on self-reported symptoms with no medical nor biological assessment routinely performed to characterize the adverse events, and information about adverse events described in this paper was sometimes incomplete because approximately 30% of participants did not answer the call of the investigators (versus 55% in the TRuST pharmacovigilance pilot [4]). Classification of adverse events remains thus debatable. Nevertheless, this follow-up represented the only feasible alternative in the study context. Moreover, the case of a participant with G6PD deficiency who was excluded by the study intervention but was later treated by the malaria program and developed hemolysis is informative: it suggests that the embeddedness between the project’s facilitators and the community allowed to detect serious events potentially related to the intervention.

### Lost in translation: Cultural aspects in the description of adverse drug reactions

When the investigator questioned participants about the symptoms reported, there was sometimes no exact match with the clinical picture described verbally and interpreted by the physician. At times, participants may not know which option between those proposed by the questionnaires best matches what they are feeling and may therefore select the closest available category; in other cases, symptom representations may differ substantially. For example, complaints described as cardiac symptoms may sometimes correspond to digestive symptoms or anxiety-related manifestations; dizziness was frequently reported in the questionnaire as dyspnea. These results indicate that to strengthen the appropriateness of community-based pharmacovigilance of antimalarial drugs, it must remain simple enough to be implemented in the field by nonmedical professionals, while also being attentive to cultural aspects and to representations of disease and medication within the study population, eventually requiring *ad hoc* social sciences study before implementation [27]. One simple solution tested by Pereira et al. [20] consists of a single screening question about whether a side effect occurred, followed by systematic contact by health staff to explore and characterize the symptom. The limitation of this approach is that it does not allow high-risk situations to be identified in advance or prioritized within the potentially substantial workload it may generate.

### Adherence to the treatment

Reported adherence (approximately 80% for 7 days primaquine) was relatively satisfactory, in comparison with findings from other studies conducted in similar cultural settings. For example, adherence reached 93.3% among participants in the 3S pilot in Amazonas [19] but lower estimates have been reported in an observational study conducted in six municipalities of the Brazilian Amazon, with 88% adherence by simple self-report and 78.2% when pill counts were taken into account [35], as well as in another Brazilian Amazon study reporting 86.4% adherence at day 7 (81.7%-90.1%) according to a strict criterion [36].

However, several limitations must be acknowledged: potential social-desirability bias, potential selection bias because participants who complied with follow-up may also have been more likely to adhere to treatment, and memory bias because tablet counts were sometimes reported during visits conducted several days later.

## Conclusions

In the context of implementing new treatment protocol against *Plasmodium vivax* based on the use of tafenoquine, health authorities might require enhanced monitoring of participants to track the safety of introducing this innovation in remote areas and among hard-to-reach populations.

This need is accentuated in the case of intensive interventions targeting the asymptomatic transmission of this parasite, such as MDA, TDA, and SeroTaT interventions using 8-aminoquinolines at curative doses, where the benefit-risk balance must be carefully monitored. The lessons learned from this study on the feasibility of such a follow-up in very complex field conditions, and on the perceived and reported tolerability of this treatment by participants in a PvTDA, aim to inform decision-makers, researchers, and caregivers and contribute to building sustainable interventions for malaria elimination. This follow-up approach may also inform patient monitoring strategies beyond malaria in settings facing similar population-level challenges.

## Acknowledgements

We would like to thank all the study population that participated in the CUREMA project. We also wish to express our sincere gratitude to all the professionals involved in the project, especially the facilitators, nurses, doctors and supervisors.

A special mention goes also to the support teams in the CHU Guyane for their contributions to data management activities as well as their regulatory and ethical support.

We would also like to acknowledge the support provided by Appsolute SAS in developing the smartphone app for the CUREMA project.

## Statements and declarations

### Ethical considerations

The CUREMA project received ethical approval from the Ethics Committee of the Ministry of Health of Suriname (CMWO 005/22, granted on June 9, 2022), and by the Oswaldo Cruz Foundation (Fiocruz) research ethics committee (CEP number: 5.210.165) and the Brazilian National Research Ethics Commission (CONEP number 5.507.24), granted on July 5, 2022. The study complies with French Law No. 2012-300 (Jardé Law) on research involving human participants, Good Clinical Practice guidelines (version dated December 15, 2016), and the principles outlined in the Declaration of Helsinki. All data were managed in accordance with the CNIL’s (Commission Nationale Informatique et Liberté) MR-001 reference methodology and the General Data Protection Regulation (GDPR).

The CUREMA project is registered on ClinicalTrials.gov ( NCT05540470).

### Consent to participate

Each participant to the CUREMA project (intervention and cross-sectional surveys) gave written consent to participation after clear and appropriate information.

### Consent for publication

Not applicable

### Data availability

The anonymized datasets generated and/or analysed during the current study are not publicly available due given the particularly sensitive nature of these data concerning a population carrying out an activity punishable by law, but are available from the corresponding author on reasonable request.

