## Supplementary table 1 and figure 1 for "Active follow-up of *Plasmodium vivax* radical treatment in a mobile and hard-to-reach population in the Amazon: results from the CUREMA project"

### Supplementary materials

Table 1: Analyse multivariée EI sous-groupe primaquine

| Inclusion site | Albina vs mean of all the sites | 1.01[0.41-2.48] | 0,004 |
| --- | --- | --- | --- |
| Inclusion site | Ampuma vs mean of all the sites | 0.23[0.09-0.62] | ** |
|  | Ilha Bela vs mean of all the sites | 0.79[0.19-3.25] |  |
|  | Oiapoque vs mean of all the sites | 2.33[0.97-5.59] | * |
|  | Paramaribo vs mean of all the sites | 0.72[0.25-2.05] |  |
|  | Upper Lawa River vs mean of all the sites | 3.19 [0.34-29.73] |  |
| Sex | Female | 1 | 0,003 |
|  | Male | 0.26[0.1-0.64] |  |
| Primaquine daily dose mg/Kg | Per additional 0.1 mg/Kg/day | 2.25[1.07-4.73] | 0,024 |

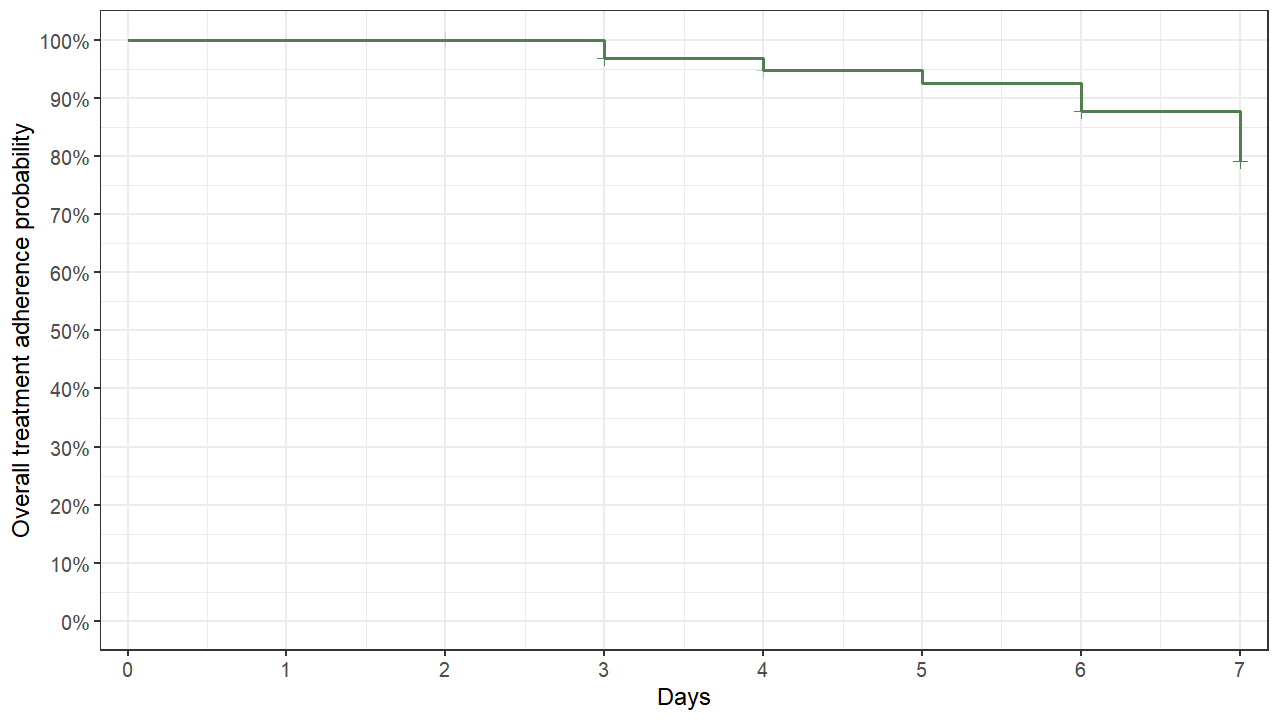

Figure 1: Kaplan-Meier illustration of the 7-day primaquine and chloroquine treatment adherence.
